# Population-based surveillance of drug-resistant tuberculosis in Southern Mozambique based on whole genome sequencing

**DOI:** 10.1101/2023.05.03.23289244

**Authors:** Carla Mariner-Llicer, Belén Saavedra Cervera, Edson Mambuque, Shilzia Munguambe, Neide Gomes, Luis Villamayor, Irving Cancino-Muñoz, Manuela Torres-Puente, Dinis Nguenha, Durval Respeito, Gustavo Tembe, Mariana G. López, Iñaki Comas, Alberto L. García-Basteiro

## Abstract

Mozambique is considered a high multidrug-resistant (MDR-TB) country by the World Health Organization (WHO) (3.7% new MDR-TB cases). The last national WHO survey was performed in 2008. Nowadays, South Africa and Eswatini are reporting an increase in MDR-TB cases heightened, in Eswatini, by the spread of a MDR-TB strain harbouring the *rpoB*_I491F mutation not detected by XpertUltra/RIF. Due to the concerning MDR-TB situation in its neighbouring countries, it is crucial to review the current resistance status in Mozambique. Our aim is to depict the current prevalence of drug-resistance to first and second-line drugs and track its trends since 2008. We analysed 312 *Mycobacterium tuberculosis* genomes from a population-based survey conducted in Manhiça in 2018 and looked for drug-resistance mutations included in the recently published WHO catalogue and compared with the prevalences reported in 2014 and 2008. We found that the overall resistance increased in 2014 (15.9%) and 2018 (14.4%) was slightly higher than the one reported in 2008. Although, new multidrug/rifampicin resistance cases remained consistent with the 2008 prevalence study (3.5%) indicating that MDR-TB is not spreading as rapidly as in neighbouring countries. Importantly, we detected a high isoniazid (INH) prevalence not associated with MDR-TB (4.2%, 5.5% and 7.6% in 2008, 2014 and 2018, p-value=0.03) suggesting that a sizeable number of cases are INH-resistant before starting treatment and not detected by XpertUltra/RIF. Fortunately, no mutations associated with second-line drug-resistance were found in the dataset. Our results show a stable drug-resistance situation in Manhiça with the need to monitor isoniazid resistance, and highlight the potential of WGS to be used in national surveys to expand our knowledge of drug-resistance prevalence throughout all Mozambican provinces.

## BACKGROUND

Mozambique is considered a high multidrug-resistant tuberculosis (MDR-TB) burden country [1]. The last drug-resistant (DR) survey conducted in 2008 by the World Health Organisation (WHO) [2] reported that around 3-4% of new TB cases were MDR (11% among previously treated) [2]. Neighbouring countries are reporting increasing rates of MDR-TB fueled by the dangerous combination of treatment failure and high transmission [3–5]. The limitations of surveys using culture-based drug-susceptibility testing (DST) and/or nucleic-amplification tests (Xpert MTB/RIF Ultra) became more evident when a MDR-TB clone, highly disseminated in Eswatini, was not detected due to a rare (but common in Eswatini) rifampicin (RIF) resistance mutation (*rpoB*_I491F) [4]. Considering the currently outdated DR estimates in Mozambique and the concerning MDR figures in its neighbouring countries, it is crucial to reassess the resistance profile in this setting. Thus, the aim of this study is to determine the current DR prevalence to first and second-line drugs by using genomic data, and to track its trends between 2008 and 2018. To do this, we identified if DR mutations listed in the new WHO mutation catalogue were present in isolates collected in a population-based study (2018) which underwent whole genome sequencing (WGS) [6].

## METHODS

Clinical samples were obtained from a population-based study conducted in Manhiça (Maputo, Mozambique) during 2018, including 312 *Mycobacterium tuberculosis* complex (MTBC) isolates (Xpatial-TB study) from new and retreatment cases. All isolates were WGS by short-read sequencing approach (Illumina) (PRJEB61426) and analysed using our published and validated pipeline (https://gitlab.com/tbgenomicsunit/ThePipeline) for mapping and variant calling steps, using as reference the MTBC inferred ancestor genome [7]. We screened for all the drug-resistance associated variants reported in the WHO catalogue for first and second-line drugs [6]. Furthermore, we performed an analysis based on expert-knowledge to find polymorphisms in candidate genes conferring resistance to second-line drugs as bedaquiline (BDQ) (*atpE, Rv0678* and *pepQ);* and delamanid (DLM) (*ddn, fgd1, fbiA, fbiB, fbiC* and *fbiD)* [8]. Additionally, we looked for frame-shift and truncations in *Rv0678* gene, as both mutation types have been recently associated with increments in the minimum inhibitory concentrations (MICs) for BDQ [9]. Prevalence of resistance and risk-ratios (RR) and their associated confidence intervals at 95% (95%CI), were measured for each antibiotic [10]. We calculated clustering at 5 SNPs distance to identify recent transmission [11] and quantified transmission events within clusters containing only resistant/susceptible strains [12]. We compared our results with previous phenotypic DR surveys conducted in Manhiça district in 2014 [13] and at national level in 2008 [2]. P-values were obtained with the Fisher-test.

## RESULTS AND DISCUSSION

Overall resistance prevalence to at least one drug, 14.4%, has not significantly changed when comparing the genomic study with the 2014 and 2008 phenotypic surveys (15.9% and 11.4%, p-value>0.05, respectively). In agreement, the DR percentage among new TB cases in the 2018 study was 13.0%, compared to 14.3% in 2014 [13] and 11.1% in 2008 [2] (p-value>0.05). Prevalence of resistance to isoniazid (INH), rifampicin (RIF), ethionamide (ETH), streptomycin (SM), pyrazinamide (PZA) and ethambutol (EMB) together with percentages of MDR are reported by case type (new or previously treated cases) in **Table1**. Despite the general stability of the DR situation over the years, our results reveal an increasing trend in new INH-R cases, 10.9%, compared to 9.2% in 2014 and 7.8% in 2008. The data shows that this increase is driven by a higher percentage of INH-R non-MDR strains (7.6% in 2018 vs 4.2% in 2008, p-value=0.03; 5.5% in 2014, p-value=0.42). This suggests that currently almost 1/10 new TB cases in Mozambique are already INH-R before starting treatment and undetected by Xpert MTB/RIF Ultra. In addition, we observed a higher proportion of adverse treatment outcomes (failure or death) in patients INH-R non-MDR (3/14, 21.4%) compared to INH and RIF susceptible (13/120, 10.8%) (treatment outcome was available only for 134/312 patients). All the ETH resistances reported were due to cross-resistance with INH. Importantly, we found no evidence related to BDQ-R and DLM-R in our setting which tallied with the limited use of these drugs in Mozambique.

**Table 1:** Prevalence of resistance in 2018, 2014 and 2008.

| Study | 2018 |  |  |  | 2014 [13] |  |  |  | 2008 [2] |  |  |  |
| --- | --- | --- | --- | --- | --- | --- | --- | --- | --- | --- | --- | --- |
| Resistance Prediction | WGS |  |  |  | Phenotypic DST |  |  |  | Phenotypic DST |  |  |  |
| Total patients | 312 |  |  |  | 276 |  |  |  | 1127 |  |  |  |
|  | New | Retreated | RR (95% CI) | p-value | New | Retreated | RR (95% CI) | p-value | New | Retreated | RR (95% CI) | p-value |
| Case type | 276 | 36 |  |  | 238 | 38 |  |  | 1102 | 25 |  |  |
| Resistant | 36 (13.0; 9.1-17) | 9 (25.0; 10.9-39.1) | 1.9 (1-3.6) | 0.050 | 34 (14.3; 10.4-19.3) | 10 (26.3; 15.0-42.1) | 1.8 (0.9-3.4) | 0.052 | 122 (11.1; 9.3-13.2) | 6 (23.3; 6.6-40.2) | 2.2 (1.1-4.4) | 0.044 |
| Susceptible | 240 (86.9; 83-90.9) | 27 (75.0; 60.9-89.2) | 0.9 (0.7-1.0) | 0.050 | 204 (85.7; 80.61-89.9) | 28 (73.7; 56.9-86.6) | 0.86 (0.71-1.04) | 0.060 | 980 (88.9; 86.8-90.7) | 19 (76.6; 59.8-93.4) | 0.8 (0.7-1.1) | 0.044 |
| **Any Resistance** |  |  |  |  |  |  |  |  |  |  |  |  |
| INH | 30 (10.9; 7.2-14.5) | 7 (19.4; 6.5-32.4) | 1.8 (0.8-3.8) | 0.130 | 22 (9.2; 6.2-13.6) | 6 (15.8; 7.4-30.4) | 1.7 (0.7-3.9) | 0.209 | 85 (7.8; 6.0-9.6) | 4 (15.0; 0-31) | 2.1 (0.8-5.2) | 0.130 |
| RIF | 9 (3.3; 1.2-5.4) | 6 (16.7; 4.5-28.8) | 5.1 (1.9-13.5) | 0.000 | 10 (4.2; 2.3-7.6) | 7 (18.4; 9.2-33.4) | 4.3 (1.8-10.8) | 0.001 | 40 (3.7; 2.4-5.1) | 5 (19.6; 4.0-35.1) | 5.5 (2.4-12.8) | 0.000 |
| ETH | 15 (5.4; 2.8-8.1) | 2 (5.6; 0-13.0) | 1.0 (0.2-4.2) | 0.970 |  |  |  |  |  |  |  |  |
| SM | 11 (4.0; 1.7-6.3) | 1 (2.8; 0-8.2) | 0.7 (0.1-5.2) | 0.720 | 7 (2.9; 1.4-5.9) | 4 (10.5; 4.2-24.1) | 3.6 (1.1-11.6) | 0.034 | 80 (7.4; 5.9-9.0) | 3 (9.8; 0.0-22.1) | 1.6 (0.6-4.9) | 0.370 |
| PZA | 3 (1.1; 0-2.3) | 0 | 0 | 0.530 | 3 (1.3; 0.3-3.6) | 0 |  |  |  |  |  |  |
| EMB | 8 (2.9; 0.9-4.9) | 1 (2.8; 0-8.2) | 0.9 (0.1-7.4) | 0.970 | 5 (2.1; 0.9-4.8) | 5 (13.2; 5.8-27.3) | 6.3 (1.9-20.6) | 0.003 | 15 (1.5; 0.6-2.4) | 2 (6.0; 0.0-15.4) | 5.9 (1.4-24.3) | 0.010 |
| **Multi Drug Resistance** |  |  |  |  |  |  |  |  |  |  |  |  |
| MDR/RR | 7 (2.5; 0.7-4.4) | 6 (16.7; 4.5-28.8) | 6.6 (2.3-18.5) | 0.000 | 10 (4.2; 2.0-7.6) | 7 (18.42; 7.7-34.3) | 4.4 (1.8-10.8) | 0.001 | 40 (3.6; 2.5-4.7) | 5 (20.0; 4.3-35.7) | 5.5 (2.4-12.8) | 0.000 |
| **Other Patterns of Resistance** |  |  |  |  |  |  |  |  |  |  |  |  |
| INH no MDR | 21 (7.6; 4.5-10.7) | 3 (8.3; 0.0-17.4) | 1.1 (0.3-3.5) | 0.880 | 13 (5.5; 2.9-9.2) | 1 (2.6; 0.1-13.8) | 0.5 (0.1-3.6) | 0.460 | 47 (4.3; 3.1-5.5) | 1 (4.0; 3.7-11.7) | 0.9 (0.1-6.5) | 0.950 |
| RIF no MDR | 2 (0.7; 0.0-0.03) | 2 (5.6; 0.0-0.2) | 7.7 (1.1-52.8) | 0.020 | 1 (0.4; 0.0-0.02) | 2 (5.3; 0.0-0.2) | 12.5 (1.2-134.8) | 0.010 | 2 (0.2; 0.0-0.01) | 2 (8.0; 0.01-0.26) | 44.1 (6.5-300.5) | 0.000 |

Data is presented as the number of cases (percentage; IC95%). Abbreviations: INH: isoniazid, RIF: rifampicin, ETH: ethionamide, SM: streptomycin, PZA: pyrazinamide, EMB: ethambutol, MDR/RR: MDR strains or mono-rifampicin strains, pre-XDR: pre-extensively drug-resistant: RR: Risk Ratio.

Compared with previous reports in the region, our estimate of MDR/RR prevalence (2.5%), is similar to the estimates published in 2014 and 2008 (3.8%, 3.7%) [2, 13] and to the one published in the WHO’s 2021 report (3.7%) [3]. Importantly, these results evidence that, fortunately, MDR/RR-TB is not spreading rapidly in this region. Due to the low number of previously treated patients recruited, we cannot compare the impact of MDR-TB in this group.

HIV status was available for 87.2% (272/312) of patients, 62.9% (171/272) of them were people living with HIV (PLHIV). Of those, 29 PLHIV also had an MTBC resistant strain to at least one drug (17.0%); and this proportion was lower in HIV negative patients (8.9%, 9/101). The low number of HIV negative patients enrolled in our study was not enough to draw conclusions on the association of HIV and DR. However, a similar and statistically-significant association between HIV and MTBC drug-resistance was found in the 2008 national survey [2].

Finally, we used a SNP pairwise distance analysis to evaluate transmission of DR strains in this setting of southern Mozambique. Overall, we identified 77 putative transmission events, 17 of them occurring between DR strains; this proportion (22.1%, 17/77) was significantly lower compared with transmission events between susceptible strains 60/78 (77.92%) (odds ratio=0.37; 95%CI 0.17-0.77; p-value=0.005). Then we focused on characterising transmission of the most prevalent mutations associated with first-line drug-resistance. *katG*_S315T, was observed in 56.8% (21/37) of INH-R strains, 12 of them (57.1%) were within 3 different clusters. *rpoB*_S450L was present in 53.3% (8/15) of RIF-R isolates, 2 of which (25%) were in transmission in the same cluster. Of note, the mutation *rpoB*_I491F, associated with RIF-R, was rare in our setting (n=1), despite being a major driver of MDR-TB in the neighbouring country Eswatini [16, 17].

These results show the most recent estimates on population-based drug-resistance prevalence in a high TB and HIV/TB burden district in Southern Mozambique. Our DR estimates in 2018 were similar to the ones obtained in 2014 [13] and 2008 [2] revealing a largely stable situation compared to neighbouring countries [4, 5]. Close monitoring of DR levels is essential to alert about a potential increase in transmission of MDR strains and guarantee that currently recommended standardised regimens for treating DR-TB continue to be effective. In this regard, we detected a statistically significant increase in isolates that were already INH-R (but not RIF-R) at the start of treatment. While much attention has been paid to RIF-R, especially among designers of molecular diagnostics, INH-R is now recognized as another major challenge for control and successful treatment outcomes [14, 15]. Thus, it is important to monitor the increasing trend of INH-R cases in a susceptible background. In summary, here we show how genomic surveillance allows us to dissect the drug-resistance situation in Mozambique including DR associated transmission patterns. This is particularly important in this setting since the neighbouring countries, Eswatini and South Africa, are reporting that many MDR-TB cases are driven by transmission [4, 5]. However, this does not seem to be the case in Manhiça, Mozambique. National representative population-based drug-resistance surveys using WGS will complement and expand our understanding of drug-resistance levels in all provinces of Mozambique and the impact on clinical outcomes to inform public policies.

## Data Availability

All data produced are available online at ENA repository. The project code is PRJEB61426 and will be publicly available after the manuscript publication

## ACKNOWLEDGMENTS

We would like to express our gratitude to Stefan Niemann (Research Center Borstel Leibniz Lung Center, Borstel, Germany) and to Alex Rosenthal (Office of Cyber Infrastructure and Computational Biology, National Institutes of Health, Rockville, Maryland, USA) from TB PORTALS for performing and funding the sequencing of the samples included in this study.

## AUTHORSHIP CONTRIBUTIONS

IC, AGB and MGL have contributed to the conception and the design of the work. EM, SM, NG, LV, ICM, MTP, DN, DDR and GT have processed the samples and acquired the data that have been analysed by BSC and CML. Interpretation of data has been made by CML, MGL, IC, AGB and BSC. CML, MGL, IC, BSC and AGB have drafted the manuscript and all authors have contributed to its revision. All authors have agreed to final approval for submission.

## ETHICAL STATEMENT

The study was approved by the National Bioethics Committee for Health of Mozambique (CNBS, Ref:369/CNBS/17) and the Internal Bioethics Committee of CISM. All methods were performed in accordance with the relevant guidelines and regulations. All study participants signed written informed consent after a verbal explanation and written information about the study was provided. For participants under 18 years of age, an informed consent was obtained from their relatives, parents or guardians.

